# Effect of continuous medical education and clinical imaging guidelines on reducing inappropriate computerized tomography utilization among children and young patients in a resource -limited settings: A before-and-after study

**DOI:** 10.1101/2024.08.16.24312127

**Authors:** Harriet Nalubega Kisembo, Richard Malumba, Ezra Kato Nsereko, Deborah Babirye, Victoria Nakalanzi, Francis Xavier Kasujja, Elsie-Kiguli Malwadde, Elizeus Rutebemberwa, Simon Kasasa, Dina Husseiny Salama, Michael Grace Kawooya

## Abstract

**Background:** Multi-Detector Computed Tomography (MDCT) has revolutionized healthcare delivery, significantly improving diagnostic accuracy and patient outcomes in various clinical settings. However, the overuse of CT examinations (CTEs), especially in resource-limited settings (RLS), poses a substantial public health challenge. Inappropriately performed CTEs, particularly among children and young adults, expose these vulnerable populations to unnecessary radiation risks, with 20%-50% of CTEs deemed inappropriate, and 10%-20% involving children. Despite the existence of evidence-based interventions like clinical imaging guidelines (CIGs) to curb this overuse, their availability and effectiveness in RLS are not well established.

**Objective:** This study aimed to determine the impact of continuous medical education (CME) and the introduction of clinical imaging guidelines (CIGs) on the appropriateness of CT utilization among children and young adults in selected hospitals in Uganda.

**Materials and Methods:** A before-and-after study design was employed to assess the effect of an intervention comprising CME and CIGs on appropriate CTE utilization. The intervention targeted healthcare providers (HCPs) across six public and private tertiary hospitals with available CT services over a 12-month period. Baseline data indicated a high prevalence of inappropriate CTEs among the target population. The proportion of CTEs performed for various body regions (head, paranasal sinuses, chest, abdomen, spine, trauma) and their appropriateness were retrospectively analyzed before and after the intervention, using the European Society of Radiology’s iGuide and pre-intervention study results as benchmarks.

**Results:** Post-intervention, the total number of CTEs performed increased by 33% (909 vs. 1210), with a 30% increase in public hospitals (300 vs. 608, p < 0.001) and a 41% increase in private-for-profit hospitals (91 vs. 238, p = 0.037). Specific increases were observed in head CTs (19%, 746 vs. 890, p < 0.0001) and contrasted studies (252%, 113 vs. 410, p < 0.0001). Conversely, CTEs for trauma decreased by 8% (499 vs. 458, p < 0.0001). Despite these changes, the overall proportion of inappropriate CTEs increased by 15% (38% vs. 44%, p < 0.001), with a 28% increase in inappropriate contrasted examinations (25% vs. 53%, p < 0.001) and a 13% increase in non-trauma cases (66% vs. 79%, p < 0.001). Notably, inappropriate CTEs for non-contrasted and trauma-related cases reduced by 28% (75% vs. 47%, p < 0.001) and 31% (34% vs. 14%, p = 0.0001), respectively.

**Conclusion:** The findings underscore the potential of CME and the adaptation of CIGs from high-resource settings to enhance the appropriateness of CT utilization in RLS. While the intervention notably reduced inappropriate trauma-related and non-contrasted CTEs, it also highlighted the complexity of achieving consistent improvements across all examination types. Further research is recommended to explore the determinants of successfully implementing CIGs in RLS, aiming to optimize CT utilization and improve patient outcomes.

## Introduction

Over the past two decades, advances in medical imaging technologies, particularly Multi-Detector Computed Tomography (MDCT), have significantly enhanced healthcare delivery and improved patient outcomes, especially in emergency situations, uncooperative patients, and complex clinical cases (1, 2, 3, 4, 5, 6, 7). Despite these benefits, the widespread and growing utilization of CT imaging has raised concerns about its appropriateness, especially given its significant contribution to the global collective dose from medical radiation exposure(5, 8, 9, 10). According to the United Nations Scientific Committee on the Effects of Atomic Radiation (UNSCEAR) 2020/2021 report, CT scans (CTs) have become the largest contributor to this collective dose(11). Alarmingly, studies indicate that 20%-50% of CT examinations (CTEs) performed globally are inappropriate(9, 10, 12, 13), with 10%-20% of these occurring among children (4, 14, 15).

Inappropriate CT examinations are those that do not align with established medical guidelines, are not medically justified, or could be replaced with safer, less costly, or more effective diagnostic methods. In resource-limited settings (RLS), inappropriate utilization of CT imaging is particularly concerning, manifesting as overuse, misuse, or underuse of the technology. Overutilization often occurs when clinicians order CT scans excessively or inappropriately, even when simpler, more cost-effective diagnostic modalities might suffice(5, 9, 16, 17, 18). Conversely, underutilization may result from logistical challenges, such as limited access to CT scanners, insufficient expertise in image interpretation, or financial constraints (15, 19).

While the health risk from a single CT scan is relatively low, the cumulative effect of increasing CT utilization is a significant public health concern, particularly in sub-Saharan Africa. Here, the population is notably young, with a median age as low as 15.1 years in some countries, yet MDCT scanners are rapidly being acquired(14, 16, 20). The decision to order a CT examination in an RLS is influenced by several factors, including healthcare infrastructure, resource availability, clinical guidelines, patient characteristics, and healthcare provider behaviors(19, 21). another key contributor to unnecessary imaging is the low level of knowledge and awareness among healthcare providers regarding radiation doses, risks associated with medical imaging, and adherence to referral criteria(20, 22, 23, 24, 25).

The potential risks of radiation-induced cancers, particularly among children and young adults, underscore the critical need for radiation dose optimization and informed decision-making in medical imaging. Young patients are more vulnerable to radiation risks due to their longer life expectancy, which provides a greater window for the development of radiation-induced cancers, the possibility of repeated examinations, and the higher sensitivity of their rapidly dividing tissues(26, 27, 28, 29). Overutilization of CT scans not only increases these risks but also strains healthcare resources, leading to overcrowded imaging departments, longer wait times, delays in patient care, and increased healthcare costs(12, 16, 30).

To address these challenges, evidence-based interventions have been developed and implemented, targeting different stages of the imaging process. These include initiatives such as patient and provider education, clinical decision support tools, and routine auditing and feedback, which have proven effective in reducing inappropriate imaging(1, 31, 32, 33, 34, 35). International organizations and professional bodies, such as the International Atomic Energy Agency (IAEA)(36, 37) and the International Commission for Radiation Protection (ICRP)(38), have recommended and supported the training of healthcare providers, implementation of clinical imaging guidelines (CIGs), and routine audits to enhance the appropriateness of medical exposures.

CIGs, developed by organizations like the American College of Radiology (ACR)(39), the Royal College of Radiology (RCR)(40), and the World Health Organization (WHO)(36), provide evidence-based recommendations to guide healthcare providers in ordering the most appropriate imaging tests. These guidelines have been shown to reduce inappropriate CTEs by 25% to 50%. However, while these guidelines offer valuable insights for high-income countries (HICs), their direct applicability in RLS may be limited due to differences in healthcare infrastructure, workforce capacity, financial resources, cultural considerations, and disease epidemiology.

In response to these challenges, a group of African experts convened in several workshops to discuss the adaptation of existing guidelines to suit the needs of RLS. The consensus was to adopt and adapt guidelines from the Royal College of Radiologists for English-speaking countries and from the French Society for French-speaking countries, given the limited resources. This study set out to evaluate the effect of continuous medical education (CME) and the implementation of adapted CIGs on the proportion of inappropriate CTEs performed among children and young patients in six hospitals in Uganda.

## Materials and Methods

### Ethical approval

Ethical approval was obtained from the Makerere University, School of Medicine Research and Ethics Committee (SOMREC), REF: #REC REF 2017-118, and the National Council for Science and Technology, REF: HS1313ES. Administrative clearance was also obtained from all the participating health facilities before the start of the study. All study procedures were in accordance with the ethical standards of the 1964 Helsinki Declaration. A waiver of consent was obtained from REC to access all patient records. To protect patient confidentiality, all identifying information was replaced with unique identification numbers.

### Study Design

This study employed a before-and-after design to evaluate the impact of continuous medical education (CME) and clinical imaging guidelines (CIGs) on the proportion of inappropriate CT examinations (CTEs) performed among children and young patients.

### Study Setting

The study was conducted in six tertiary public and private hospitals in Uganda. These hospitals were purposively selected based on the following criteria: national and regional referral status, university teaching hospital affiliation, and regional representation, availability of functional CT services during the study period, the ability to identify eligible cases retrospectively, and geographical representation of CT services across the country. The hospitals were categorized into three groups according to Ministry of Health service standards: two public (PH), two private-for-profit (PFP), and two private not-for-profit (PNFP) hospitals.

### Intervention

Following a baseline study that revealed high levels of inappropriate CT examinations among children and young patients(41), an intervention was introduced. This intervention comprised continuous medical education (CME) and the implementation of clinical imaging guidelines from the European Society of Radiology (ESR)-iGuide. The intervention targeted healthcare providers (HCPs), including interns, medical officers, residents, specialists, nurses, radiographers, radiologists, information technologists, and administrative staff at the participating hospitals.

The CME sessions were delivered by two radiologists (a senior consultant and a professor), a senior imaging technologist, and a clinical epidemiologist. Each session included a one-hour lecture on the basic principles of radiation protection, justification and optimization of medical exposures, radiation doses from common imaging procedures, equivalent to number of chest x-rays and risks of radiations as in Table 1 below:

**Table 1:**
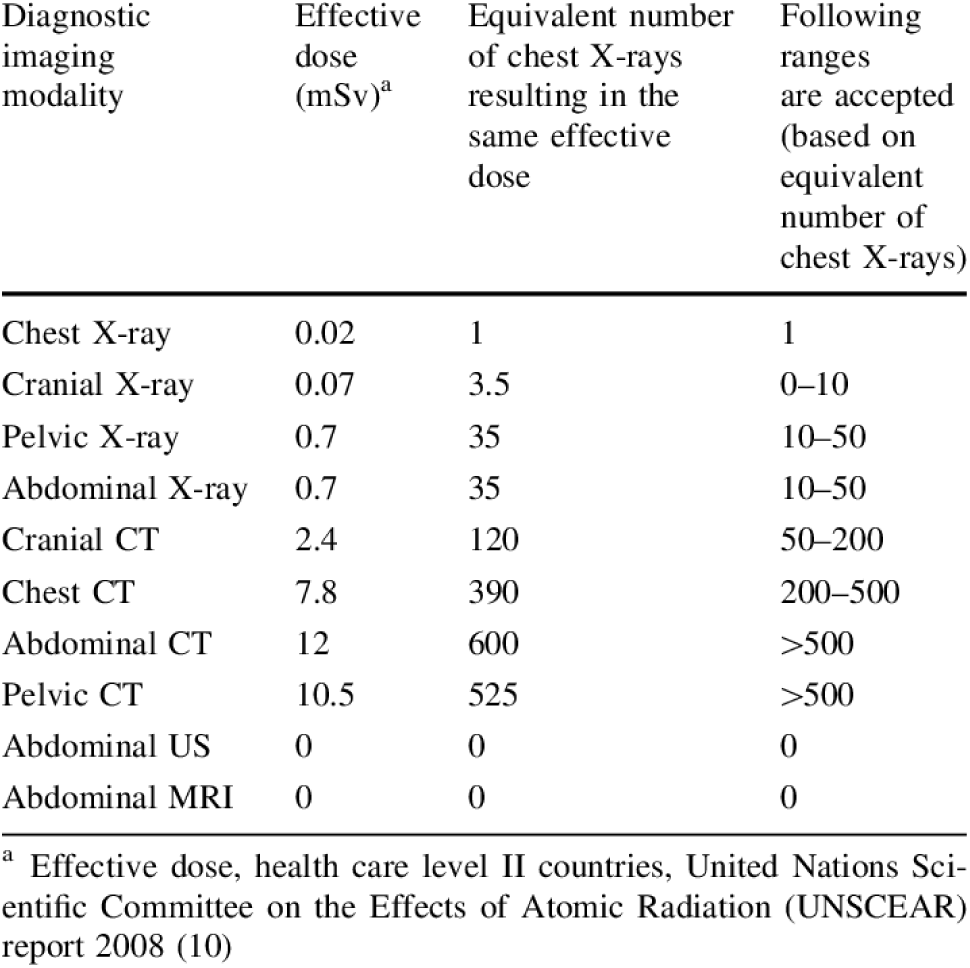
effective radiation doses for common diagnostic imaging modalities and equivalent number of chest x-rays doses.

| Diagnostic imaging modality | Effective dose (mSv)<sup>a</sup> | Equivalent number of chest X-rays resulting in the same effective dose | Following ranges are accepted (based on equivalent number of chest X-rays) |
| --- | --- | --- | --- |
| Chest X-ray | 0.02 | 1 | 1 |
| Cranial X-ray | 0.07 | 3.5 | 0–10 |
| Pelvic X-ray | 0.7 | 35 | 10–50 |
| Abdominal X-ray | 0.7 | 35 | 10–50 |
| Cranial CT | 2.4 | 120 | 50–200 |
| Chest CT | 7.8 | 390 | 200–500 |
| Abdominal CT | 12 | 600 | >500 |
| Pelvic CT | 10.5 | 525 | >500 |
| Abdominal US | 0 | 0 | 0 |
| Abdominal MRI | 0 | 0 | 0 |

<sup>a</sup> Effective dose, health care level II countries, United Nations Scientific Committee on the Effects of Atomic Radiation (UNSCEAR) report 2008 (10)

Alternative imaging modalities that use less or no ionizing radiation were also highlighted. Following the lecture, a practical interactive session was conducted, during which participants accessed the online ESR-iGuide, registered, and downloaded the app on their devices. Participants were also provided with various educational materials, including handout summaries, overheads, IAEA posters on radiation protection during CT scans, and justification of CT examinations, which were displayed in workstations.

The CME sessions were repeated 6 to 12 months after the initial training in all hospitals. A pre- and post-intervention test using a structured questionnaire was administered to participants to assess their knowledge and awareness of radiation safety and justification of medical exposures. The results have been published elsewhere. Attendance was voluntary, though the sessions also counted towards routine continuous professional development, a mandatory requirement for the renewal of HCPs’ annual practice licenses.

### Data Management and Analysis

#### Hypothesis

The null hypothesis (Ho) posited that there would be no difference in the proportion of inappropriate CT requisitions for patients aged 35 years and below before and after the intervention. The alternative hypothesis (Ha) proposed that the use of CIGs by CT prescribers would reduce inappropriate CT requisitions for common examinations in this age group by 10-15% across the six selected hospitals.

### Data Source

A decline in the use of CT scans was anticipated following the intervention, though expected to be lower than in the implementation studies (18, 42). For a hypothesized absolute decrease of 15% in CT utilization, with 90% power and a one-tailed α of 0.05, from a baseline level of 80% for patients aged 35 years and below who had CT scans performed during the study period, a minimum sample size of 148 CT request forms (CTRFs) per phase (before and after the intervention) was required to achieve a power of 0.90 and a one-tailed α of 0.05.

Retrospective retrieval from hospital archives included all consecutive eligible CTRFs for head, paranasal sinuses (PNS), chest, abdomen, spine, or trauma CTs performed from July 1 to December 31, 2020. The upper age limit of 35 years was selected as the radiation-induced lifetime attributable risk of cancer mortality from low-dose radiation plateaus beyond this age.

The principal author (HNK) screened all retrieved CTRFs, excluding unreadable records, duplicates, canceled examinations, electronic medical requests, and requests from external referrers. The analysis methods mirrored those used in the baseline study by Kisembo et al(41).

### Clinical imaging guidelines

Permission and virtual training on the use of the web-based guideline were provided by the ESR-ACR iGuide Project of the European Board of Radiology. Each requisition was independently reviewed by at least two researchers (HNK and RM) to assess the match between the clinical information provided and the clinical condition options reported by the “Consult Appropriate Use Criteria (AUC)” online computer program(43). The AUC is based on the Appropriateness Criteria developed by the American College of Radiology and embedded into the ESR-ACR iGuide, a computerized decision support (CDS) platform. All researchers had adequate experience in analyzing the appropriateness of medical tests. Disagreements were resolved through discussion and consensus. If necessary, a third reviewer (MGK) served as the tie-breaker.

### Rating of Appropriateness of CT Request Forms (CTRFs)

The clinical information on the majority of CTRFs was converted into a coding system to derive ratings of appropriateness. Each eligible requisition was manually coded into the software by entering patient demographics (age and sex) and relevant clinical indications, symptoms, diagnoses, or differential diagnoses that could justify the CTE. The computer system scored each requisition using a 1 to 9 ordinal-point rating scale, with 1 being the least appropriate and 9 being the most appropriate. Scores were categorized as follows: 7-9 indicated the most appropriate imaging study to be performed first (usually appropriate), 4-6 indicated a specialized investigation not initially indicated (may be appropriate), and 1-3 indicated that ordering should be avoided, with consideration of specialist referral or radiology consultation (not appropriate).

For this study, the 9-point rating scale was simplified into a two-grade scale, as used by Vilar-Palop et al.. A score of 7–9 was classified as appropriate, while a score of 1-6 was classified as inappropriate. If the CTRF lacked sufficient clinical information to understand the patient’s condition and evaluate the appropriateness of the examination according to the guideline, it was categorized as not justified and included in the inappropriate group. Requests that could not be categorized based on the guideline were excluded from the study. CTRFs requesting scans for multiple body parts (e.g., CT chest and abdomen) were treated as separate requests, and criteria/guidelines were applied individually to each.

### Statistical Methods

Frequency distributions and cross-tabulations were used to describe the data. The primary outcome measures were the difference in the total number of CTEs and the proportion of inappropriate CTEs before and after the intervention. These were calculated separately and compared using Pearson’s chi-square test or Fisher’s exact test, with a significance level of P < 0.05. Data was analyzed in relation to hospital category, patient age, sex, anatomical region, indication, and use of contrast media.

## Results

A total of 2,119 computed tomography examinations (CTEs) were performed on patients aged ≤ 35 years across the six participating hospitals during the two study periods. Of these, 38% (799/2,119) were females, and 23% (539/2,119) were children (aged < 18 years). The majority of the CTEs were head scans, constituting 77% (1,638/2,119) of the total. Non-contrast CTEs accounted for 69% (1,463/2,119), and trauma-related CTEs represented 45% (957/2,119).

Following the intervention, the overall number of CTEs performed increased by 33%, from 909 before the intervention to 1,210 after the intervention. This increase was particularly significant in public hospitals (PHs) and private-for-profit hospitals (PFPs), where the number of CTEs rose by 30% (300 vs. 608, p < 0.001) and 41% (91 vs. 238, p = 0.037), respectively. In contrast, private not-for-profit hospitals (PNFPs) experienced a 27% decrease in inappropriate CTEs after the intervention (518 vs. 364, p < 0.001).

Specifically, head CTEs increased by 19% (746 vs. 890, p < 0.0001), and contrasted CTEs surged by 252% (113 vs. 410, p < 0.0001) following the intervention. Conversely, CTEs performed for trauma decreased by 8% (499 vs. 458, p < 0.0001). These changes in the number of CT examinations performed for patients aged ≤ 35 years before and after the intervention are detailed in Table 2.

**Table 2:** The total number (n) and changes in the numbers of different CT examinations performed on patients aged ≤ 35 years before and after the intervention at the six participating hospitals.

|  |  | after-<br>intervention<br>N (%) | % Change<br>before and after<br>intervene | P-values | 95% Confidence<br>Intervals |
| --- | --- | --- | --- | --- | --- |
| n | 909 (43) | 1,210 (57) | 14.2 | <0.001 | (0.0994, 0.1846) |
| **Hospital** |  |  |  |  |  |
| A | 58 (6) | 197 (16) | 9.9 | 0.056 | (0.0177, 0.1803) |
| B | 193 (21) | 231 (19) | -2.14 | 0.584 | (-0.0982, 0.0554) |
| C | 33 (4) | 41 (3) | -0.24 | 0.830 | (-0.2429, 0.1949) |
| D | 107 (12) | 377 (31) | 19.39 | <0.001 | (0.1170, 0.2708) |
| E | 103 (11) | 100 (8) | -3.07 | 0.462 | (-0.1123, 0.0509) |
| F | 415 (46) | 264 (22) | -23.83 | <0.001 | (-0.3074, -0.1692) |
| Hospital<br>Category |  |  |  |  |  |
| Public Hospital | 300 (33) | 608 (50) | 17.25 | <0.001 | (0.1061, 0.2389) |
| Private for<br>Hospital | 91 (10) | 238 (20) | 9.66 | 0.037 | (0.0169, 0.1763) |
| Private not for<br>profit | 518 (57) | 364 (30) | -26.91 | <0.001 | (-0.3326, -<br>0.2056) |
| **Sex** |  |  |  |  |  |
| Male | 572 (63) | 741(61) | 1.69 | 0.532 | (-0.0698, 0.0360) |
| Female | 331 (37)) | 468 (39) | 2.19 | 0.529 | (-0.0462, 0.900) |
| Missing | 6 (10) | 1 (0) | -0.58 |  |  |
| **Age categories** |  |  |  |  |  |
| < 1 | 36 (4) | 61(5) | 1.08 | 0.807 | (-0.0733, 0.0949) |
| 1 <= 5 | 63 (7) | 80 (7) | 0.32 | 0.940 | (-0.0862, 0.0798) |
| 6 <= 10 | 48 (5) | 66 (6) | 0.17 | 0.968 | (-0.0820, 0.0854) |
| 11 <= 15 | 52 (6) | 55 (5) | -1.17 | 0.784 | (-0.0955, 0.0721) |
| 16 <= 20 | 115(13) | 161(13) | 0.66 | 0.872 | (-0.0737, 0.0869) |
| Above 20 | 595(66) | 786 (65) | -0.5 | 0.847 | (-0.0557, 0.6928) |
| Missing | 0 | 1 (0) | 0.08 |  |  |
| **Age categories** |  |  |  |  |  |
| <18 | 232 (26) | 307 (25) | -0.15 | 0.968 | (-0.0758, 0.0728) |
| ≥18 | 677 (75) | 902 (75) | 0.07 | 0.975 | (-0.0427, 0.0441) |
| Missing | 0 (0) | 1 (0) | 0 |  |  |
| **Anatomic Region** |  |  |  |  |  |
| None | 34 (4) | 2 (0) | -3.57 | 0.791 | (-0.1213, 0.0499) |
| Head | 746 (82) | 890 (74) | -8.52 | <0.001 | (-0.1252, -0.0452) |
| spine | 16(2) | 61 (5) | 3.28 | 0.567 | (-0.0518, 0.1174) |
| Abdomen | 48 (5) | 101 (8) | 3.07 | 0.502 | (-0.0552, 0.1138) |
| Chest | 34 (4) | 134 (11) | 7.33 | 0.195 | (-0.0097, 0.1563) |
| PNS | 20 (2) | 12 (1) | -1.21 | 0.800 | (-0.0974, 0.0732) |
| Others | 11 (1) | 10 (1) | -0.38 | 0.931 | (-0.0895, 0.0819) |
| **Contrast** |  |  |  |  |  |
| Not contrasted | 796 (88) | 800 (66) | -21.45 | 0.931 | (-0.0895, 0.0819) |
| Contrasted | 113 (12) | 410(34) | 21.45 | <0.001 | (0.1383, 0.2907) |
| **Trauma** |  |  |  |  |  |
| Non-trauma | 412 (45) | 702 (58) | 12.7 | <0.001 | (0.0666, 0.1874) |
| Trauma | 497 (55) | 458 (38) | -16.83 | <0.001 | (-0.2307, -0.1059) |
| Missing | 0 (0) | 50 (4) | 4.13 |  |  |

The overall proportion of inappropriate CTEs significantly increased by 15%, from 38% before the intervention to 53% after the intervention (p < 0.001). Notably, inappropriate contrasted and non-traumatic CTEs increased by 28% (25% vs. 53%, p < 0.001) and 13% (66% vs. 79%, p < 0.001), respectively. However, there was a reduction in the proportion of inappropriate non-contrasted and trauma-related CTEs by 28% (75% vs. 47%, p < 0.001) and 21% (34% vs. 14%, p < 0.001), respectively.

The number, proportion, and difference in proportions of inappropriate CTEs out of the total number of cases analyzed in 2018 and 2020 for patients are summarized in Table 3.

**Table 3.** The number (n), proportion (%) and the difference in proportions of inappropriate CT examinations out of the total number of the cases analyzed before and after intervention for patients aged ≤ 35 years.

|  | Baseline | Post-intervention | % change | p-value | 95% CI |
| --- | --- | --- | --- | --- | --- |
| **Inappropriateness** | **347 (38)** | **645 (53)** | **+15.14** | **<0.001** | **(0.2382, 0.3626)** |
| **Sex** |  |  |  |  |  |
| Male | 139 (40) | 287 (44) | 4.44 | 0.386 | (-0.5530, 0.1441) |
| Female | 207 (60) | 358 (56) | -4.15 | 0.337 | (10.1259, 0.4286) |
| **Age categories** |  |  |  |  |  |
| < 1 | 13 (4) | 56(9) | 4.93 | 0.549 | (-0.0776, 0.1762) |
| 1 ≤ 5 | 27 (8) | 45 (7) | -0.8 | 0.899 | (-0.1335, 0.1175) |
| 6 ≤ 10 | 24 (7) | 37 (6) | -1.18 | 0.852 | (-0.1380, 0.1144) |
| 11 ≤ 15 | 26 (8) | 37 (6) | -1.75 | 0.781 | (-0.1434, 0.1084) |
| 16 ≤ 20 | 52 (15) | 92 (14) | 14.26 | 0.905 | (-0.1278, 0.1132) |
| Above 20 | 205 (59) | 377 (59) | -0.63 | 0.883 | (-0.0900, 0.0774) |
| **Age categories 2** |  |  |  |  |  |
| <18 | 105 (30) | 200 (31) | 0.75 | 0.893 | (-0.1013, 0.1163) |
| $\geq 18$ | 242 (70) | 444 (69) | -0.9 | 0.807 | (-0.0816, 0.0632) |

| **Anatomic Region** |  |  |  |  |  |
| --- | --- | --- | --- | --- | --- |
| None | 34 (10) | 2 (0.3) | -9.49 | 0.653 | (-0.2211, 0.0313) |
| Head | 239 (69) | 412 (64) | -5 | 0.195 | (-0.1248, 0.2481) |
| spine | 7 (2) | 38 (6) | 3.87 | 0.674 | (-0.0896, 0.1670) |
| Abdomen | 27 (8) | 92 (14) | 6.48 | 0.375 | (-0.0589, 0.1887) |
| Chest | 25 (7) | 87 (14) | 6.29 | 0.395 | (-0.0613, 0.1871) |
| PNS | 8 (2) | 7 (1) | -1.22 | 0.852 | (-0.1416, 0.1172) |
| Others | 7 (2) | 7 (1) | -0.93 | 0.888 | (-0.1388, 0.1202) |
| **Contrast** |  |  | 0 |  |  |
| Not contrasted | 259 (75) | 302 (47) | -27.82 | <0.001 | (0.3555, -0.2009) |
| Contrasted | 88 (25) | 343 (53) | 27.82 | <0.001 | (-0.1731, 0.3833) |
| **Trauma** |  |  | 0 |  |  |
| Non-trauma | 228 (66) | 507 (79) | 12.89 | <0.001 | (0.0569, 0.2001) |
| Trauma | 119 (34) | 88 (14) | -20.65 | 0.001 | (-0.3179, -0.0951) |

NB; \*p-value <0.05. NA = Assumptions for the normal approximation to the binomial were not met. The percentages shown above are for inappropriateness

## Discussion

This study aimed to evaluate the impact of continuous medical education (CME) and clinical imaging guidelines (CIGs) on reducing inappropriate computed tomography examinations (CTEs) in a resource-limited setting (RLS). CME is well recognized for its role in enhancing the quality of medical practice. Integrating radiation protection and the application of CIGs into these sessions was intended to improve the appropriateness of CTE requisitions(35, 44, 45, 46).

Our findings reveal a 33% overall increase in the number of CT examinations performed, accompanied by a 15% rise in inappropriate CTEs. This suggests that the intervention did not optimally change clinical practice behavior regarding the use of CT examinations.

There are could be several explanations for these findings. First, the increase in CTEs might reflect a broader global trend of heightened use of cross-sectional imaging, particularly among young patients. Factors contributing to this trend include advancements in CT technology, greater availability of CT scanners, increased public awareness and demand for advanced diagnostic imaging, and changes in disease epidemiology, such as the rising burden of non-communicable diseases (3, 4, 6, 15, 16, 21, 28, 29, 47, 48, 49).

Second is the ineffectiveness of CIGs, which were originally developed for high-income countries (HICs), in altering CT utilization behaviors in an RLS, aligns with the existing literature on guideline implementation (19, 21, 42, 50). For these guidelines to be effective in an RLS, they must be adapted to local contexts, with consideration for constraints, resources, and collaboration among stakeholders.

Third, since a previous study from the same setting reported low utilization of advanced imaging modalities like CTs, the intervention might have inadvertently raised awareness among healthcare workers about the availability and utility of CT scans, leading to an overall increase in their requisition, even when inappropriate(51).

The intervention’s effects varied across hospital types. While public and PFP hospitals saw significant increases in CTEs by 30% and 41%, respectively, PNFP hospitals experienced 27 % decrease in CTEs. This decline in PNFPs might be attributed to prior similar interventions that reinforced compliance and increased awareness among imaging referrers. This aligns with findings from Forsetlund et al., (44) who noted that multi-session educational meetings are more effective than single sessions for knowledge retention and behavior change.

Trauma related CT utilization, which accounted for 45% of all CTEs, significantly decreased by 20% after the intervention. The reduction in inappropriate trauma-related CTEs by 16% may be partly due to the national lockdown and travel restrictions during the COVID-19 pandemic in 2020(52). While the overall number of accidents may have decreased, the severity of injuries might have increased, justifying the need for CT examinations for high-energy injuries. The intervention likely provided evidence-based criteria that supported more judicious decision-making by imaging referrers when ordering CTEs.

Traumatic brain injuries, a significant contributor to inappropriate CTEs, remain a priority area for interventions aimed at reducing unnecessary scans. A study by Babirye et al. demonstrated that a significant proportion of head CTEs for mild traumatic brain injuries could have been avoided by applying clinical decision rules like the Canadian CT Head Rule (CCHR) (53).

The 28% increase in inappropriate contrasted CTEs could be due to the lack of alternative imaging modalities like ultrasound or MRI, which do not use ionizing radiation but are often unavailable in RLS (18, 19, 21, 48, 50, 54).

We however also observed a lack of radiologist consultation by IRs before ordering CT examinations, contributing to the escalation of inappropriate CT utilization(38, 55, 56). Radiologists play a crucial role as gatekeepers, ensuring that patient exposures are clinically justified by approving appropriate requisitions and rejecting those that are not. Studies have shown that IRs are more cautious in their ordering practices when they know their requests will be scrutinized and possibly rejected(57, 58).

### Limitations

Several potential biases could affect the estimated impact of the intervention, including secular trends, seasonal effects, the duration of the intervention, and random fluctuations. This underscores the need for robust experimental or quasi-experimental designs to evaluate strategies for reducing inappropriate imaging utilization in RLS. Additionally, the limited number of CME sessions may have constrained the intervention’s impact, although this was enhanced by an interactive learning model incorporating lectures, case discussions, and hands-on practice. The study’s short follow-up period raises uncertainty about whether similar effects would be observed over more extended periods. Moreover, the impact of various intervention components was not measured separately, making it likely that certain interventions had a more significant influence on referral practices than others. However, the multi-center representation of CT services in this study increases the generalizability of the results to hospitals with CT services across Uganda. Other limitations noted in the baseline study also apply to this analysis.

## Data Availability

currently, Data will be available upon request from the corresponding author, later deposition will be in Institutional Makerere University repository

## Acknowledgments

We acknowledge the administrative officers, CT in-charges, CT imaging technologists, research assistants, and radiologists from the participating hospitals for their invaluable support throughout this study. We extend our gratitude to all healthcare workers who actively participated in the CME sessions. Special appreciation goes to the ESR-ACR iGuide Project secretariat of the European Board of Radiology for their unwavering support in terms of training and for allowing free access to their guidelines. We also thank the IAEA Radiation Protection of Patients unit for providing free educational and training resources.

## Conclusion and Implications

One of the most significant findings to emerge from this study is that CME for HCPs and the adoption of CIGs from HICs have the potential to improve the appropriateness of CT utilization in RLS, particularly by reducing inappropriate trauma-related and non-contrasted CTEs among children and young patients.

However, further research is needed to focus on the determinants of implementing CIGs from HICs into RLS, with the goal of improving evidence based clinical practice behaviors. In these environments, a phased approach is recommended, focusing on creating awareness, encouraging dialogue, and adapting existing CIGs to local resources. This can lay the groundwork for more comprehensive, locally tailored quality improvement processes in the future.

